# Pan-cancer Graph-based Cancer Detection Using the Cell-free DNA Methylome

**DOI:** 10.64898/2026.08.26.26361432

**Authors:** Lin Zhao, Yong Zeng, Dor D. Abelman, Wenjun Lin, Ping Luo

## Abstract

**Motivation:** Cell-free DNA methylation provides a minimally invasive signal for early cancer detection and tissue-of-origin prediction. Most methods represent methylation measurements as independent fixed-window features and therefore do not explicitly model relationships among genomic regions.

**Results:** We developed PANGEM (Pan-cancer Graph-based Cancer Detection Using the Cell-free DNA Methylome), a graph-learning framework that represents genomic bins as nodes and integrates CpG context, genomic proximity, and sample-specific methylation similarity in the graph topology. Across five repeated stratified train-test splits, PANGEM achieved the highest mean performance among evaluated methods, with an AUROC/AUPR of 0.997±0.003/1.000±0.000 for binary cancer detection and macro-AUROC/AUPR of 0.977±0.027/0.870±0.086 for multiclass tissue-of-origin prediction. In the independent INSPIRE cohort, 72 of 78 cancer cases (92.3%) exceeded the binary classification threshold, and PANGEM correctly classified 9 of 17 head and neck cancer cases (52.9%), the highest accuracy among evaluated methods. Subnetwork analysis further identified recurrent, graph-connected methylation patterns, including a 111-DMR subnetwork with increased methylation in cancer samples.

**Availability and Implementation:** Source code for this study is available at https://github.com/GenTensor/PANGEM. Contact

**Supplementary information:** Supplementary data are available at Bioinformatics online.

## Introduction

Cell-free DNA (cfDNA) consists of short DNA fragments released into body fluids, primarily during apoptosis or necrosis (3). It supports non-invasive cancer detection and monitoring (13), including applications investigated in clinical studies and FDA-approved diagnostic assays (11; 6; 2). Among liquid-biopsy signals, cfDNA methylation is particularly informative because aberrant methylation patterns can indicate both tumor presence and tissue of origin (15).

Most cfDNA methylation analyses represent samples as high-dimensional vectors of fixed genomic windows (bins) and apply statistical feature selection followed by conventional machine learning models (4; 14; 16). This representation treats genomic regions as independent features, although CpG sites in islands, shores, and shelves often exhibit coordinated methylation and nearby genomic bins can behave as coherent units (9). Models must therefore infer these relationships indirectly, which may increase sample requirements and limit biological interpretation. Fixed-bin analyses may also overlook broader differentially methylation regions (DMRs) of variable genomic length (12). Graph neural networks (GNNs) provide a natural way to encode such relationships and learn context-dependent representations through neighborhood aggregation (10).

We developed PANGEM, a graph-based framework that represents each cfDNA methylation profile as a sample-specific graph. Genomic methylation bins form the nodes, with node features combining methylation measurements and local genomic context. Edges integrate a static prior based on CpG annotation and genomic proximity with dynamic similarity derived from each sample’s methylation profile. This hybrid construction preserves shared genomic structure while allowing connectivity to reflect patient-specific epigenetic variation, avoiding the rigidity of fully static graphs and the noise and computational cost of constructing unconstrained dynamic graphs. A dual-branch, identity-aware, edge-gated GNN then learns graph-level representations for cancer detection and cancer-type classification. The graph representation also enables subnetwork-level interpretation by identifying connected discriminative methylation regions that are not readily captured by conventional methods. The complete workflow is summarized in Figure 1. To our knowledge, PANGEM is the first framework to represent genome-wide cfDNA methylation profiles as graphs for cancer prediction.

**Figure 1.**
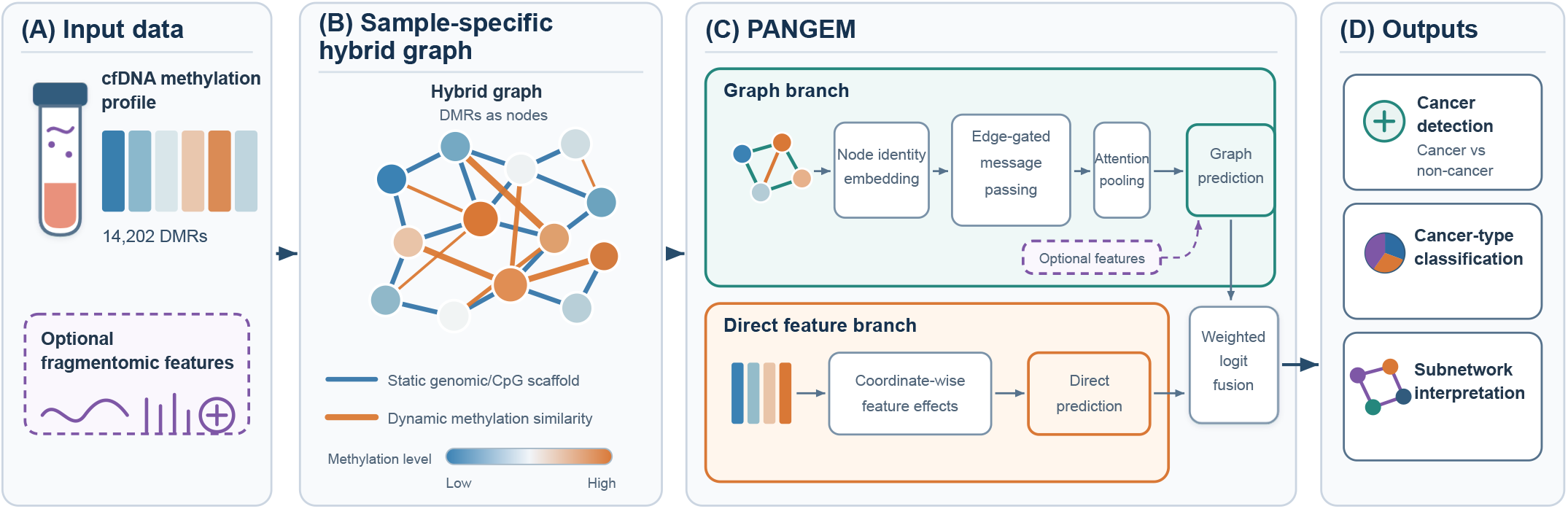
Overview of the PANGEM workflow. (A) cfDNA methylation profiles across 14,202 DMRs provide the primary input, with fragmentomic features included optionally. (B) Each sample is represented as a hybrid graph that integrates a static scaffold based on genomic proximity and CpG context with sample-specific methylation similarities. (C) PANGEM integrates an edge-gated graph branch with a direct feature branch; optional fragmentomic features enter the graph-level representation before weighted logit fusion. (D) The framework supports cancer detection, cancer-type classification, and subnetwork interpretation.

## Materials and Methods

### Dataset and Preprocessing

In this study, we used the processed cell-free methylated DNA immunoprecipitation sequencing (cfMeDIP-seq) dataset generated by Zeng et al. (19). In their study, sequencing reads were processed using MEDIPIPE v1.1.0 (20), and methylation signals were quantified in consecutive 300-bp genomic bins. Quality control, normalization, batch-effect correction, and differential methylation analysis were performed as previously described. We used the resulting normalized methylation matrix and published pan-cancer DMR panel as input. This panel comprised 14,202 hypermethylated DMRs.

The dataset comprised 1,074 cfMeDIP-seq samples collected from nine independent cohorts. For model development, we used the baseline cohort consisting of 468 paired-end cfMeDIP-seq samples. An additional 78 samples from the independent INSPIRE cohort (14) were reserved exclusively for external validation and were not used during model training or hyperparameter optimization. Because the INSPIRE cohort contains several cancer types absent from the development cohort, it provides a stringent evaluation of the model’s ability to generalize to previously unseen cancer types.

The development cohort included healthy controls, patients with cancer, and individuals with Li-Fraumeni syndrome (LFS), including survivors, previvors, and LFS-positive individuals with cancer (Supplementary Table S1). Because cancer-negative LFS profiles differ from those of healthy controls (15), survivors and previvors were excluded from binary classification but retained for multiclass classification. In addition, healthy controls were excluded from multiclass classification.

### Graph Construction

Each cfMeDIP-seq profile was represented as a sample-specific graph *G*^(*s*)^ = (*V, ε*^(*s*)^, **X**^(*s*)^, **E**^(*s*)^). The shared node set contained the 14,202 bins, **X**^(*s*)^ ∈ ℝ^*N ×*10^ contained methylation and genomic-context features, and 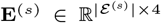 contained edge attributes. A shared genomic scaffold defined candidate edges, which were reweighted using each sample’s methylation profile to obtain the final hybrid graph. Unless otherwise stated, all graph-construction and model-capacity parameters were selected through the internal cross-validation procedure described in Section 2.4, rather than specified a priori. The complete search spaces and selected configurations are reported in Supplementary Table S2.

#### Node Feature Extraction

For bin *v*_*i*_, let 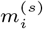, *c*_*i*_, and chr_*i*_ denote its normalized methylation value, genomic centre, and chromosome. We omit the sample superscript from the node-feature expressions below. Bins were ordered within each chromosome. The chromosome-restricted sets

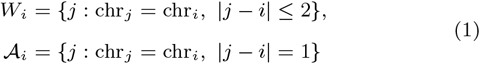

defined a five-bin local window and the available immediately adjacent bins. We then computed

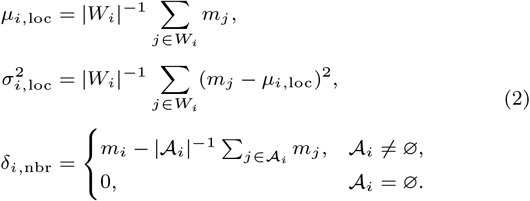

Windows were truncated rather than allowed to cross chromosome boundaries. Additional four indicators encoded island, shore, shelf, or open-sea context, and two sinusoidal terms encoded chromosome-normalized position, with 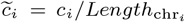 . Defining slog(*x*) = sign(*x*) log(1 + |*x*|), the resulting feature vector was

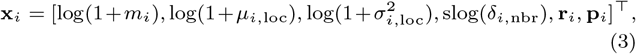

where **r**_*i*_ ∈ {0, 1}^4^ and 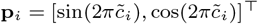 .

#### Static Prior Graph

To construct the static graph, we assigned CpG-context weights *w*_*i*_ of 1.0, 0.8, 0.6, and 0.4 to islands, shores, shelves, and open sea, respectively. These values encoded a monotonic heuristic prior over CpG context rather than calibrated biological effect sizes. The shared static score combined the geometric mean of the endpoint weights with genomic proximity:

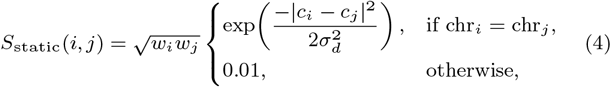

with *σ*_*d*_ = 1000 bp. After min-max normalization, the 15 highest-scoring neighbours per node formed the candidate scaffold. Restricting subsequent sample-specific comparisons to this sparse scaffold avoided unconstrained all-pairs dynamic comparisons while retaining genomically informed candidates.

#### Dynamic Edge Reweighting and Fusion

After obtaining the static graph, for each sample, let **z**_*i*_ ∈ ℝ^4^ denote the first four, methylation-derived components of **x**_*i*_. These components were standardized across all 14,202 nodes to obtain 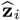 . Components with negligible variance were centered without rescaling. Dynamic similarity was then evaluated only for edges in the static candidate scaffold:

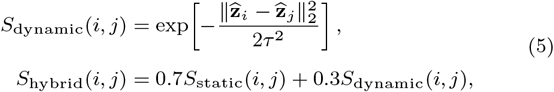

where *τ* = 1. The eight highest-scoring candidates per node were retained, and the union of these selections was symmetrized. For each retained edge, *a*_*ij*_ = |*m*_*i*_ − *m*_*j*_ | was min-max scaled over the sample’s retained edges to obtain 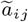. It was set to zero when all retained edges had the same value. The final edge attribute was

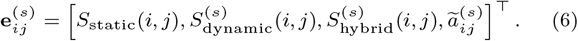

Thus, each retained edge was represented by its static, dynamic, and hybrid similarity scores and its scaled methylation difference.

### Dual-Branch Identity-Aware Edge-Gated GNN

Each sample was processed by two branches. The graph branch learned neighborhood-dependent representations through edge-gated message passing, whereas the direct feature branch operated on the node-feature matrix without neighborhood aggregation and retained coordinate-wise feature effects. Both branches used the shared node indexing across samples.

For the graph branch, each node *v* had a learnable identity embedding 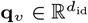, which was concatenated with its feature vector and projected to the initial hidden state:

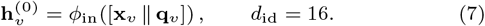

Here, ∥ denotes vector concatenation, and *ϕ*_in_ consists of a linear layer, batch normalization, ReLU, and dropout. The resulting hidden state had 64 dimensions. We omit the sample superscript from the graph-branch equations below.

For each edge (*u, v*), the four edge attributes were encoded and used to gate the message from *u* to *v*:

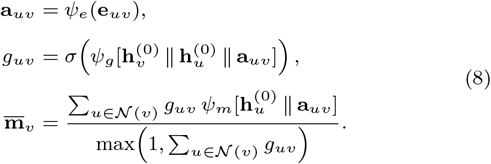

Here, *ψ*_*e*_ is the edge-attribute encoder, *ψ*_*m*_ is the message network, *ψ*_*g*_ is the gating network, and *σ* is the logistic sigmoid. The set *N*(*v*) contains the neighbors of node *v*, and the denominator normalizes the aggregate by the total gate mass. An update network *ψ*_*u*_ combined the aggregate with the target-node state. A residual connection retained part of the initial representation:

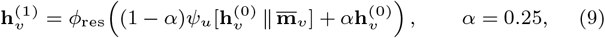

where *ϕ*_res_ applies batch normalization, ReLU, and dropout. The final model used one message-passing layer. Attention-based global pooling aggregated the final node states into a graph representation 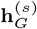, and a linear classifier produced the graph-branch logits

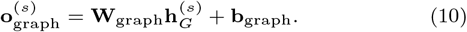

Here, **W**_graph_ and **b**_graph_ are the classifier weight matrix and bias vector. The pooling operation is described in Supplementary Methods.

The direct feature branch assigned class-specific weights to every combination of node and feature. Let *N* = 14,202 denote the number of nodes, *F* = 10 the node-feature dimension, and *C* the number of classes. With **A** ∈ ℝ^*N ×F ×C*^, its logit for class *c* was

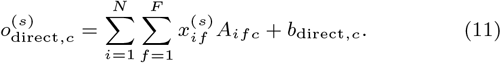

Here, 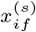 is feature *f* of node *i, A*_*ifc*_ is its learned class-specific weight, and *b*_direct,*c*_ is the corresponding bias. Writing the class logits as 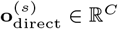, the two branches were combined at the logit level:

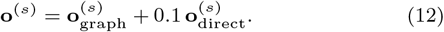

The direct-branch coefficient was tuned and chosen as 0.1. A softmax produced class probabilities, and the network was trained end-to-end using inverse-frequency class-weighted cross-entropy, with *C* = 2 for binary detection and *C* = 8 for multiclass classification.

The primary PANGEM model used only the methylation-derived graph inputs. A separate supplementary analysis (Supplementary Table S4.) evaluated whether adding fragmentomic features improved performance, as they were previously shown to provide predictive value in the study by Zeng et al.

### Model Evaluation

PANGEM was evaluated for binary cancer detection and multiclass cancer type prediction. Performance was assessed using the area under the receiver operating characteristic curve (AUROC) and the area under the precision-recall curve (AUPR), with macro-averaging applied for multiclass evaluation.

We compared PANGEM with the machine-learning methods evaluated by Zeng et al. In their approach, principal component analysis (PCA) was applied to the methylation matrix, and the resulting principal components were used as input features for seven classifiers: random forest (RF), gradient boosting (GB), *k*-nearest neighbors (KNN), logistic regression (LR), support vector machine (SVM), eXtreme Gradient Boosting (XGBoost), and linear discriminant analysis (LDA). All baseline hyperparameters were set according to the configurations reported by Zeng et al.

We used five repeated stratified 90%/10% train-test splits. To assess robustness to sampling variation rather than performance under split-specific optimization, we selected hyperparameters once by 5-fold cross-validation within the first 90% training subset and held them fixed across all subsequent train-test splits. Every method used identical split assignments, and results are reported as mean ± standard deviation across held-out test sets. Search spaces and selected values are reported in Supplementary Table S2.

## Results

### Evaluation on Held-out Samples

PANGEM achieved the highest performance for binary cancer detection (Figure 2), with an AUROC of 0.997 ± 0.003 and an AUPR of 1.000± *<* 0.001. LR was the strongest baseline, achieving 0.992 ± 0.012 and 0.999 ± 0.002, respectively. Although all methods effectively distinguished cancer from healthy samples, PANGEM showed the highest mean performance with low variability across splits.

**Figure 2.**
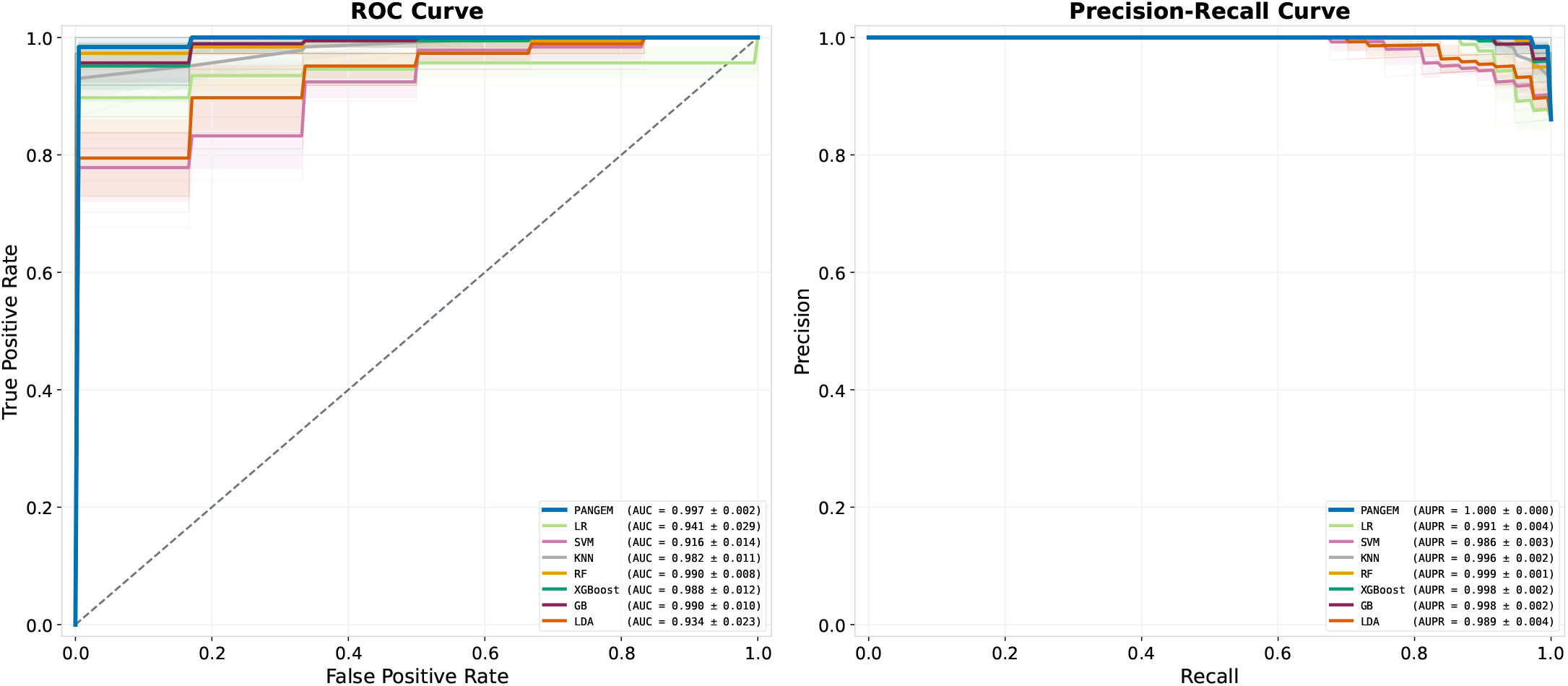
Cancer-versus-healthy classification. ROC (left) and PR (right) curves compare PANGEM with seven baseline models across five repeated 90%/10% splits. Legends report mean area under the curve *±* standard deviation.

For multiclass classification, PANGEM also achieved the highest mean performance (Figure 3), with a macro-AUROC of 0.977 ± 0.027 and a macro-AUPR of 0.870 ± 0.086. Thus, PANGEM consistently achieved the highest mean AUROC and AUPR across both classification tasks.

**Figure 3.**
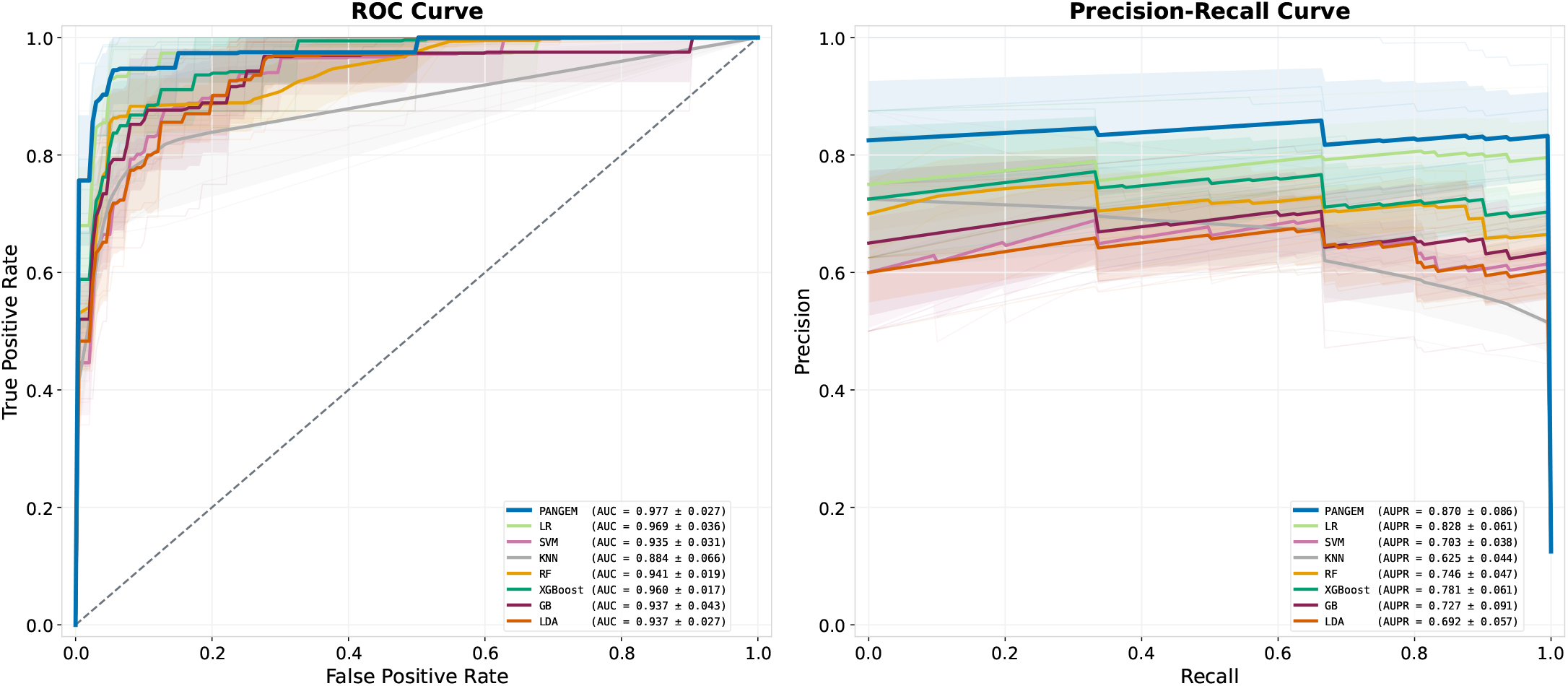
Multiclass pan-cancer classification. Macro-averaged one-versus-rest ROC (left) and PR (right) curves compare PANGEM with seven baselines across five repeated 90%/10% splits.

### Evaluation on INSPIRE Cohort

For external evaluation, five PANGEM models were trained on the full binary-eligible development set and their probabilities averaged for each of 78 INSPIRE patients. In the binary analysis, 72 samples (92.3%) exceeded the 0.5 threshold (Figure 4). Rates were 95.0% for breast, 94.1% for head and neck (HNC), 100.0% for mixed, and 89.5% for ovarian cancer. Their median scores ranged from 0.999 to 1.000. Melanoma transferred less strongly, with a median of 0.604 and 7 of 9 positive predictions. The absence of healthy controls precluded assessment of discrimination or specificity.

**Figure 4.**
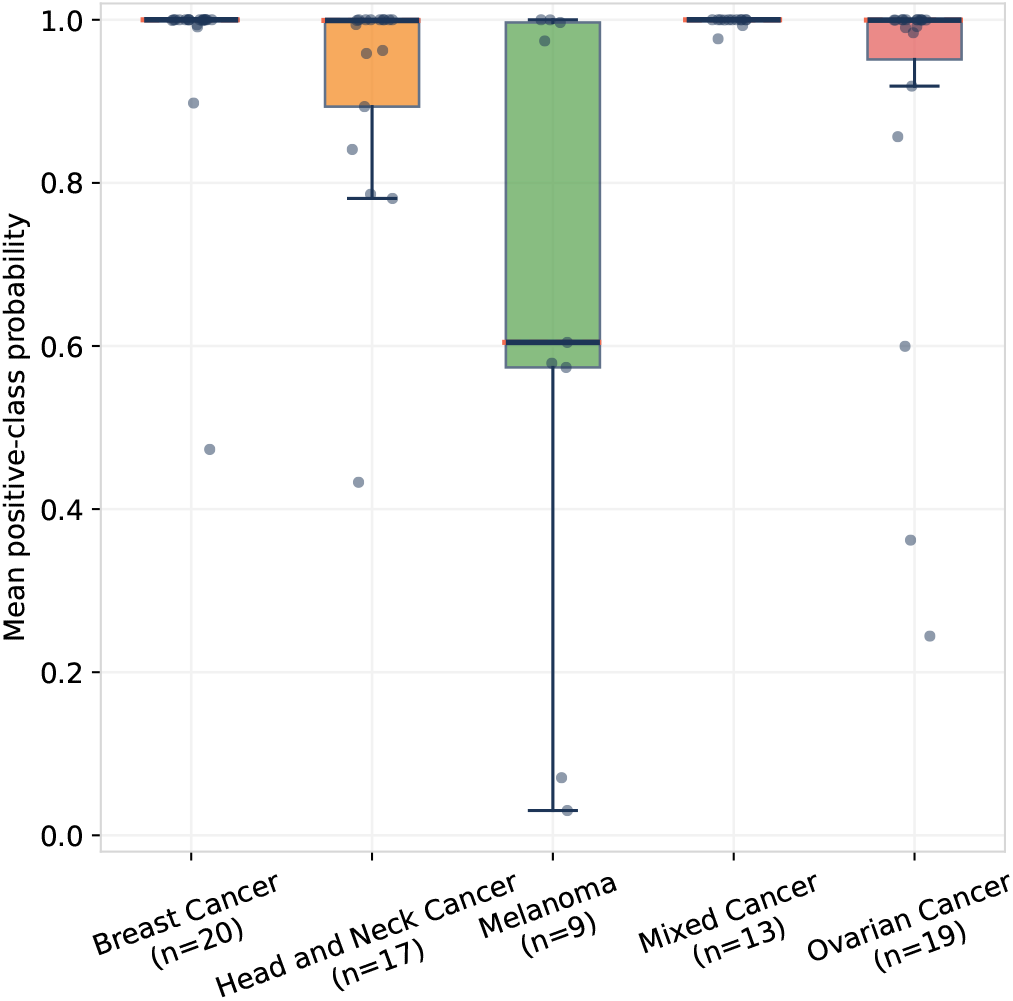
PANGEM cancer type probabilities in INSPIRE. Boxplots show patient-level scores by cancer type, averaged across five random seeds. Because INSPIRE contained no healthy controls, the analysis assesses positive-class transfer rather than cancer-versus-healthy discrimination.

Multiclass transfer was evaluated using the 17 HNC samples, the only cancer type shared between the INSPIRE and training cohorts. Class probabilities from PANGEM were averaged across five models trained on the full development cohort. PANGEM correctly classified 9 samples (52.9%), achieving the highest accuracy and substantially outperforming five of the seven baselines, supporting improved transfer of tissue-of-origin predictions to the independent INSPIRE cohort (Supplementary Figure S1).

### Subnetwork Analysis

To determine whether PANGEM prioritized biologically interpretable DMR structures, we mapped model-selected nodes to their bins and assessed recurrence across five training seeds. Direct class contributions were converted to one-versus-rest margins, and connected high-scoring bins were grouped using edges present in at least 50% of relevant sample graphs. Calls recurring in at least three seeds were considered stable, and adjacent bins were merged into regions. Multiclass interpretation was restricted to brain, prostate, lung, and HNC because these groups had sufficient samples.

In the binary model, 280 recurrent nodes formed 224 regions and 74 subnetworks (Supplementary Figure S3). The largest contained 111 bins across 17 chromosomes (Figure 5) and had higher mean methylation in cancer than healthy samples (4.81 versus 1.05; *P* = 1.90×10^*−*26^; *d* = 0.66). All 111 bins had panel-wide FDR *<* 0.05, but only 10 ranked among the top 1,000 individual effects (Supplementary Figure S2), indicating a distributed signal. Other recurrent regions mapped near *SIX1, HOXD3*, and *IGF2BP1*, which have reported roles in colorectal cancer, prostate-cancer grade or recurrence, and lung adenocarcinoma, respectively (18; 5; 17).

**Figure 5.**
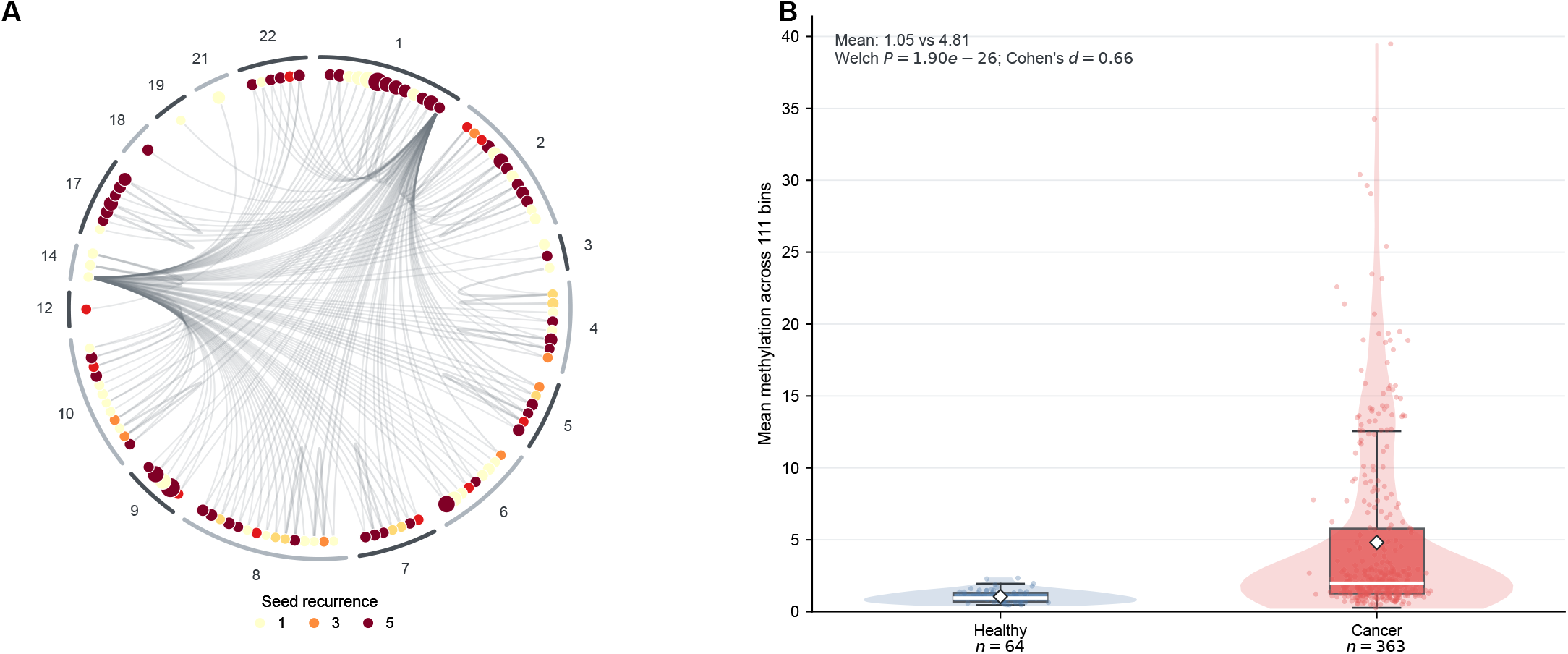
Largest stable binary PANGEM subnetwork. (A) Topology of 111 bins across 17 chromosomes; color shows five-seed recurrence and size shows median direct contribution. (B) Mean methylation was higher in cancer than healthy samples (4.81 versus 1.05; *P* = 1.90 *×* 10^*−*26^; *d* = 0.66).

In the multiclass model, 1,120 stable nodes formed 958 regions and 206 cancer-type-associated subnetworks (Supplementary Figure S4). Brain-associated regions mapped near *MIR137HG* and *TBX3* (7; 1). HNC-associated modules included *SOX1* and *HOXC6*, with *SOX1* located within a 21-bin module and previously reported to exhibit oncogenic activity in glioblastoma (8). Lung subnetworks contained a recurrent region near *IGF2BP1*, which is overexpressed in lung adenocarcinoma and promotes malignant phenotypes (17) (Supplementary Figure S3), while prostate subnetworks again included *HOXD3*.

Stable nodes showed no promoter or CpG-island enrichment beyond the regulatory composition of the input panel (Supplementary Figure S5). Binary odds ratios were 0.87 for promoters, 0.94 for CpG islands, and multiclass values were likewise close to one. Thus, recurrence and connectivity, rather than annotation enrichment, identified the coordinated methylation patterns.

### Ablation Study

To examine how graph construction affected predictive performance, we conducted an ablation study and evaluated four PANGEM configurations on the same five held-out splits, without retuning hyperparameters (Supplementary Tables S3). The combined model retained the direct methylation and hybrid graph branches. Three graph-only variants used hybrid, static-only, or dynamic-only graph construction.

For binary detection, the combined model achieved near-ceiling performance, with an AUROC of 0.997 and an AUPR of 1.000. The hybrid graph also retained strong predictive performance, reaching an AUROC of 0.944 and an AUPR of 0.991. Its mean AUROC exceeded the static-only and dynamic-only variants by 0.085 and 0.029, respectively. The margin between the combined and graph-only models supports retaining the direct methylation pathway alongside the graph representation in the full architecture. For multiclass classification, the combined model achieved a macro-AUROC of 0.977 and a macro-AUPR of 0.870. Among the graph-only variants, the hybrid graph achieved the highest mean macro-AUROC at 0.726 and improved both mean metrics relative to the static-only and dynamic-only constructions.

Overall, the ablation results support the full PANGEM architecture and the use of hybrid graph construction. Combining static genomic context with sample-specific methylation relationships gave the strongest mean performance among the biologically structured graph-only variants, while the integrated model achieved the best results for both tasks. Because all variants used the combined model’s hyperparameters, these comparisons assess robustness under a common training configuration rather than the maxima of separately optimized models.

## Discussion

PANGEM combines a static genomic prior with sample-specific methylation relationships to model cfDNA methylomes as graphs. The model achieved the highest performance for both cancer-versus-healthy detection and multiclass classification. Ablation results supported the hybrid topology, which improved the graph branch over single-component controls. On the independent INSPIRE cohort, PANGEM produced cancer-positive predictions across five diagnostic groups, including types absent during development. However, the lack of healthy controls prevents this cohort from providing a complete external assessment of discrimination or specificity.

The subnetwork analysis identified recurrent, graph-connected methylation patterns within the predefined DMR panel. The largest binary subnetwork captured a coordinated cancer-associated signal across genomic regions, although few constituent bins ranked among the strongest individual effects. This suggests that graph-based analysis can organize lower-ranked DMRs into coordinated modules that may be overlooked by locus-wise ranking alone. Recurrent regions mapped near cancer-related genes, and stable cancer-type-associated subnetworks were identified for brain, prostate, lung, and HNC, although proximity-based gene assignments do not establish regulatory relationships.

This study has several limitations. The development cohort was relatively small and class-imbalanced, and the DMR panel was derived from the full baseline cohort, potentially introducing selection bias and optimistic internal performance estimates. Although all methods used the same panel, allowing controlled model comparisons, this shared bias remains. The independent INSPIRE cohort was not involved in DMR selection but lacked healthy controls, limiting assessment of specificity and overall discrimination. Future studies should evaluate PANGEM in larger prospective cohorts using independently derived or training-specific DMR panels and experimentally validate the biological relevance of the identified methylation subnetworks.

Together, these findings demonstrate the value of sample-specific graph construction for cfDNA methylation analysis. By integrating shared genomic priors with patient-specific relationships, PANGEM achieved strong performance for cancer detection while identifying recurrent methylation subnetworks that are not captured by conventional approaches. These results highlight graph-based modeling as a promising framework for integrating genome-wide methylation information and improving the interpretability of cfDNA-based cancer detection.

## Data Availability

All data produced in the present study are available upon reasonable request to the authors.

https://doi.org/10.5281/zenodo.15191455

## Data Availability

All data produced in the present study are available upon reasonable request to the authors.

https://doi.org/10.5281/zenodo.15191455

## Data availability

The data underlying this article were originally published by Zeng et al. and is publicly available in Zenodo at https://doi.org/10.5281/zenodo.15191455.

## Acknowledgments

This research was enabled in part by support, software, and computational resources provided by SHARCNET, SciNet, Compute Ontario, Calcul Québec, and the Digital Research Alliance of Canada (alliancecan.ca). Specifically, calculations were performed using the Nibi, Trillium, and Rorqual clusters.

