## Supplementary Material for "Pan-cancer Graph-based Cancer Detection Using the Cell-free DNA Methylome"

#### Supplementary Methods

##### Graph Readout and Fragmentomic Feature Extension

The main text defines the one-layer identity-residual edge-gated update. Restoring the sample superscript omitted from those equations, let $\mathbf{h}_v^{(1,s)}$ denote the final node state for node $v$ in sample $s$. Attention-based global pooling assigns each node the weight

$$\eta_v^{(s)} = \frac{\exp\Big(\rho(\mathbf{h}_v^{(1,s)})\Big)}{\sum_{j\in\mathcal{V}} \exp\Big(\rho(\mathbf{h}_j^{(1,s)})\Big)}, \tag{1}$$

where $\rho$ is a learnable scalar attention-score network. A learnable feature-transformation network $\zeta$ then produces the graph embedding

$$\mathbf{h}_G^{(s)} = \sum_{v\in\mathcal{V}} \eta_v^{(s)}\,\zeta(\mathbf{h}_v^{(1,s)}). \tag{2}$$

The primary PANGEM model used this methylation-derived graph embedding directly, giving graph-branch logits

$$\mathbf{o}_{\text{graph}}^{(s)} = \mathbf{W}_{\text{graph}}\mathbf{h}_G^{(s)} + \mathbf{b}_{\text{graph}}. \tag{3}$$

Motivated by the performance gains reported by Zeng et al., we additionally incorporated fragmentomic profiles, fragment-ratio statistics, nucleosome-distance distributions, and motif proportions into PANGEM. These features were concatenated with the graph-level representation before classification. For comparison with the primary model, the fragmentomic feature extension used the same five repeated stratified 90%/10% splits and identical split assignments. Results are summarized in Supplementary Table S4.

#### Supplementary Figures

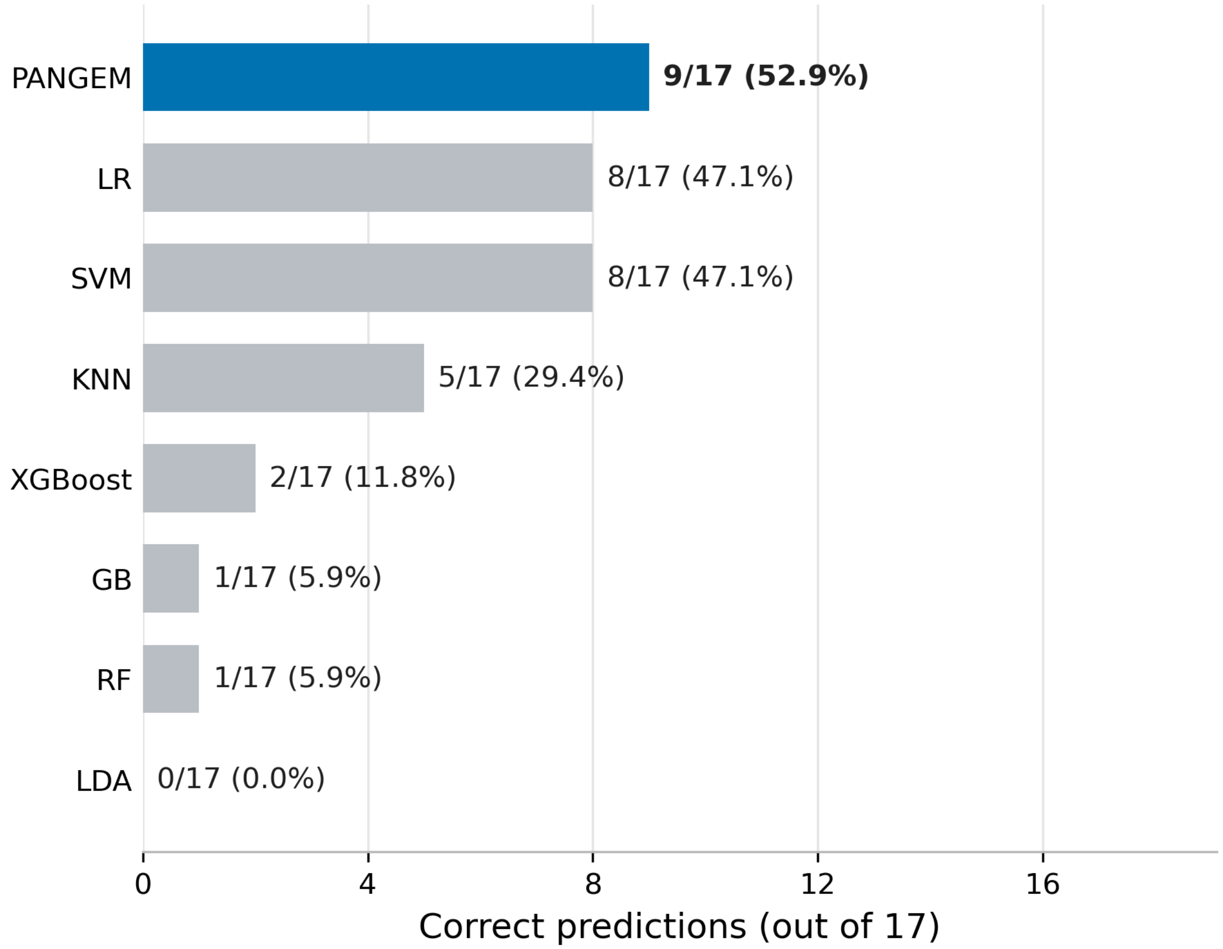


Figure S1: **Multiclass classification of head and neck cancer in the independent INSPIRE cohort.** Bars report the number and percentage of 17 head and neck cancer samples assigned to the correct cancer type by PANGEM and seven input-matched conventional machine-learning models. All methods used the same 14,202 methylation DMRs without sample-level auxiliary features. For each method, class probabilities were averaged across models trained with seeds 42, 43, 44, 45, and 46. PANGEM correctly classified 9 of 17 samples (52.9%); LR and SVM, the strongest baselines, each classified 8 of 17 (47.1%).

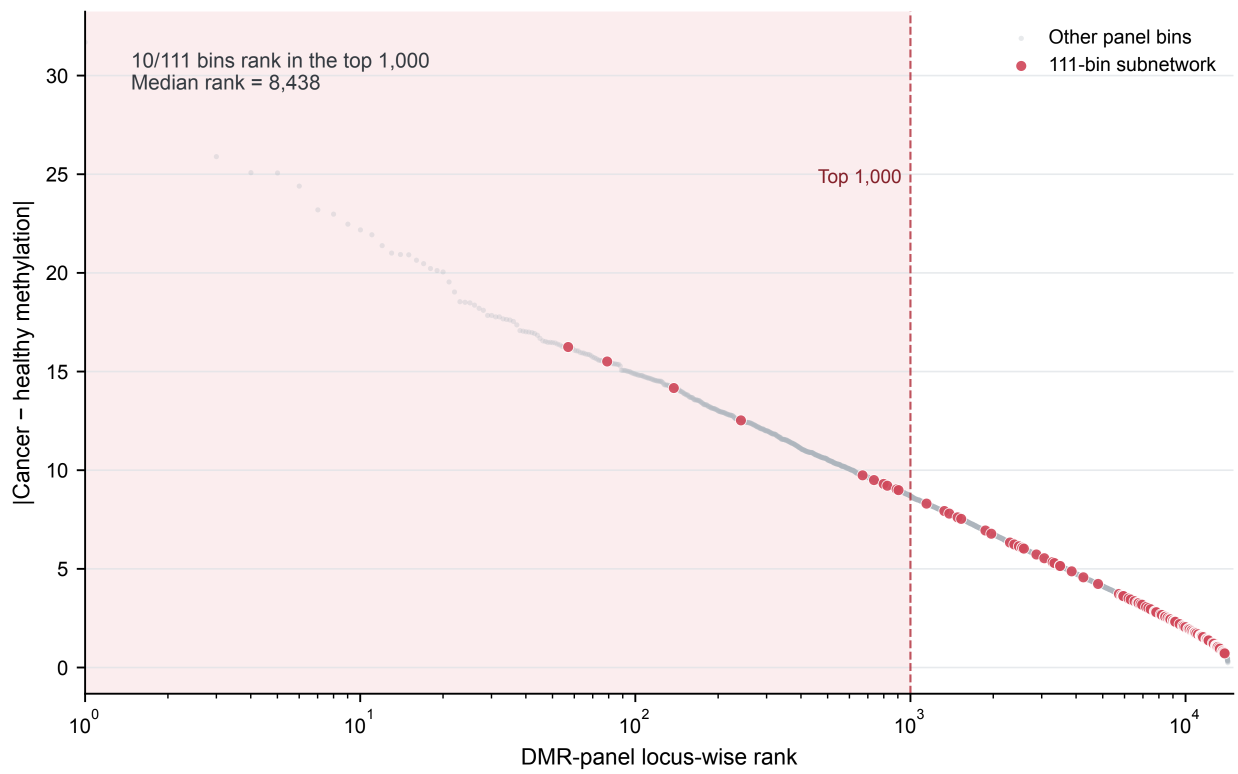


Figure S2: **Locus-wise ranking of DMRs in the largest stable binary PANGEM subnetwork.** The 111 subnetwork bins were distributed across the locus-wise ranking of the 14,202-bin DMR panel. Ten bins ranked among the top 1,000 individual effects, and the median rank was 8,438.

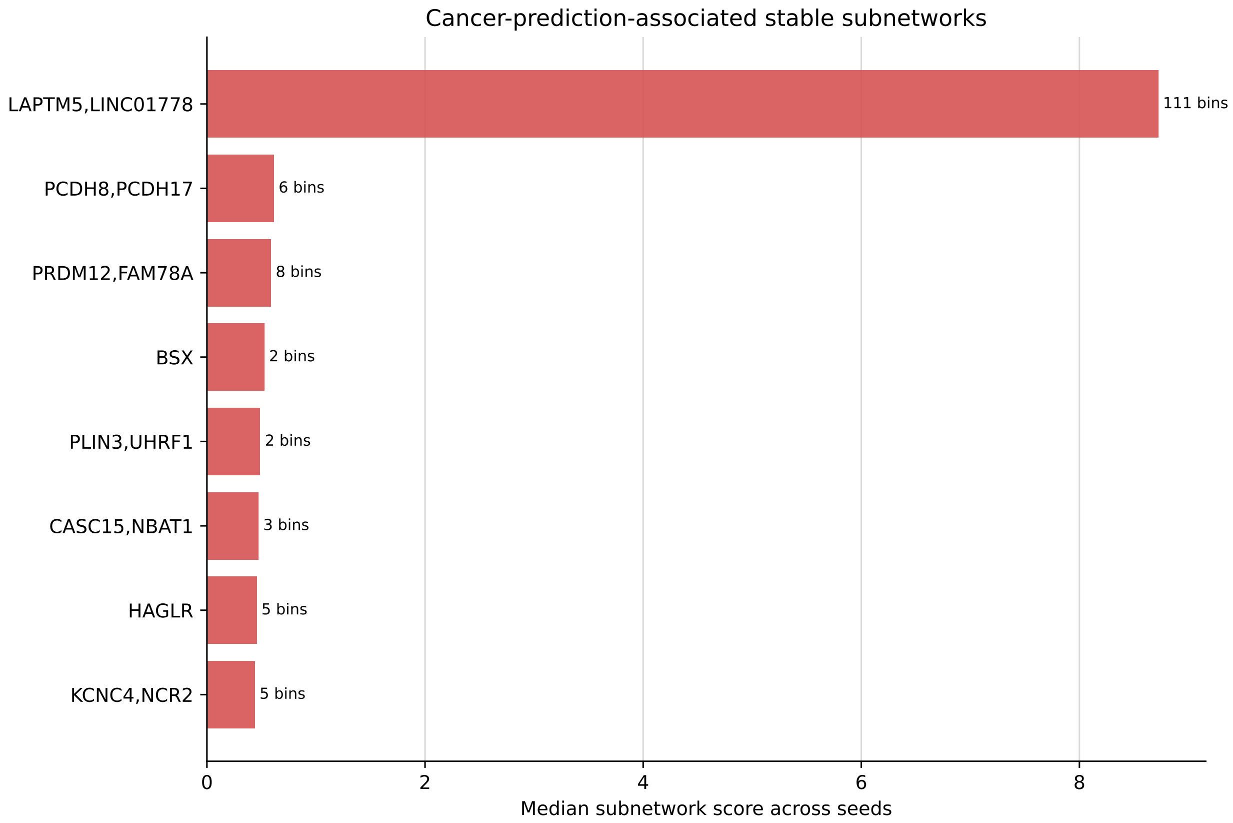


Figure S3: **Stable subnetworks associated with binary cancer prediction.** Bars show the median direct-contribution score across five trained seeds. Gene labels indicate the nearest annotated gene.

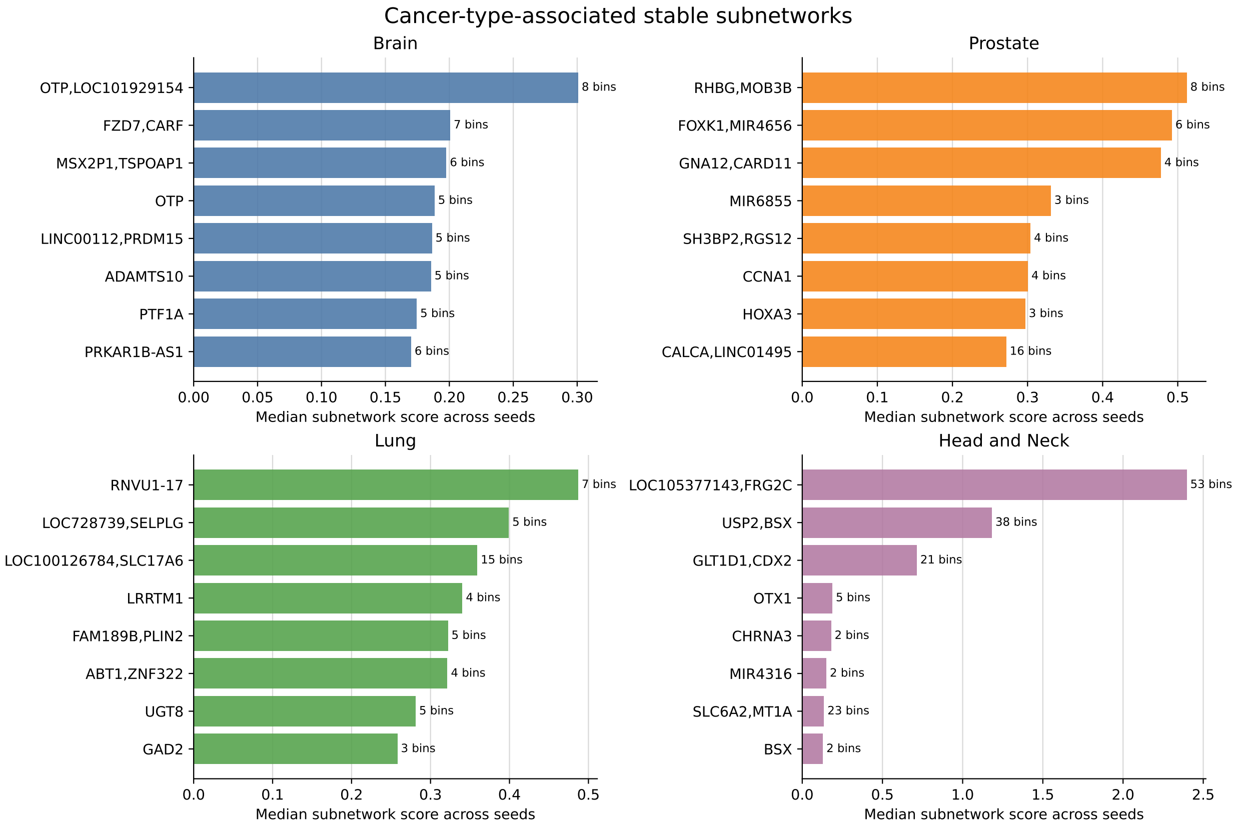


Figure S4: **Cancer-type-associated stable subnetworks in the multiclass model.** For each of the four adequately powered cancer types, bars show the top recurrent subnetworks ranked by median direct residual-head score across five seeds. Bar labels indicate the number of DMR bins in each subnetwork.

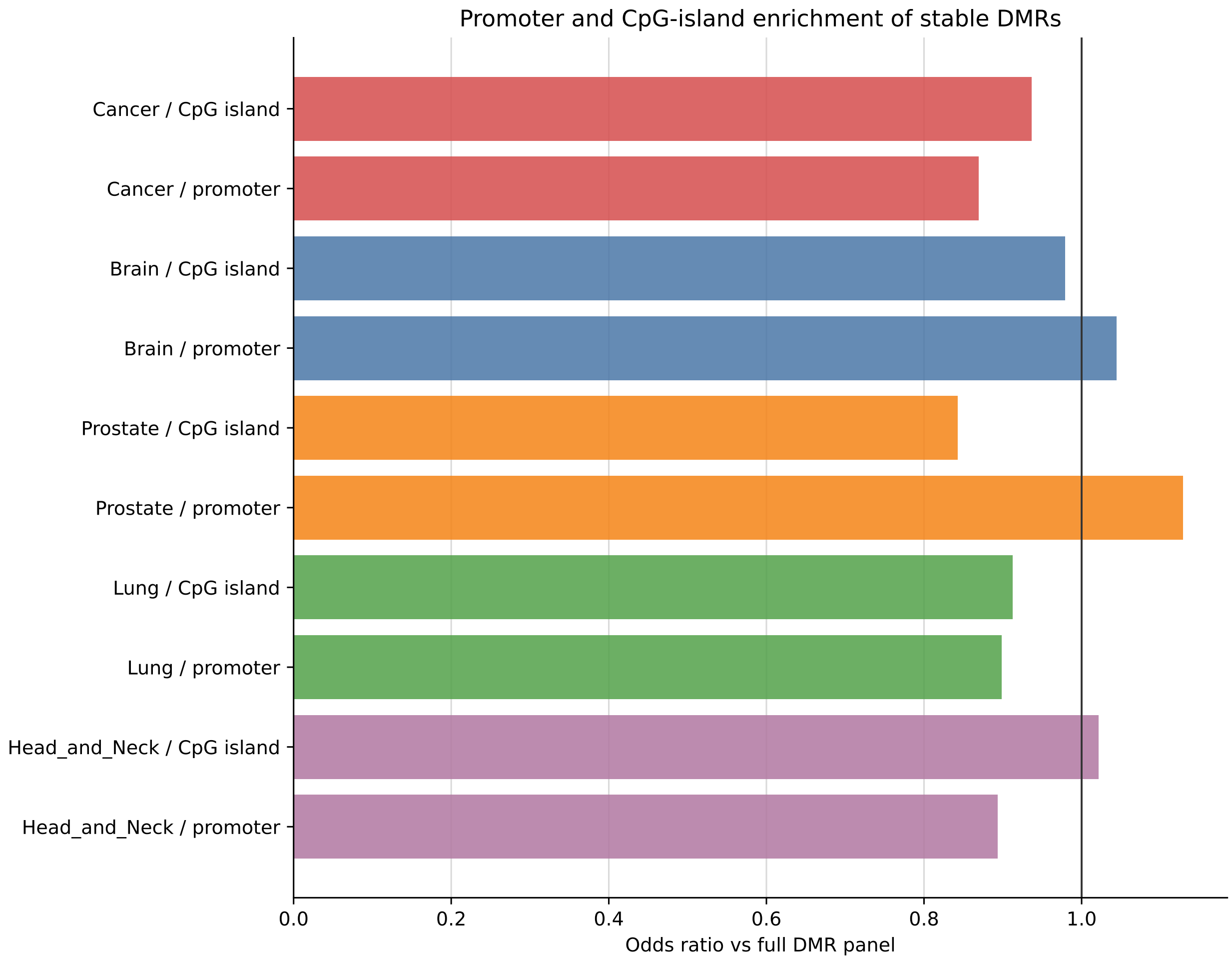


Figure S5: **Promoter and CpG-island enrichment of stable model-selected DMRs relative to the full DMR panel.** Odds ratios were computed against the 14,202-bin DMR panel background rather than against the whole genome. Values near one indicate that the stable model-selected bins largely reflect the regulatory composition of the input panel.

### Supplementary Tables

Table S1: **Composition of the dataset.**

| Diagnostic group | Samples, $n$ |
|---|---|
| *Baseline cohort* | |
| Healthy controls | 64 |
| Brain cancer | 152 |
| Prostate cancer | 97 |
| Lung cancer | 59 |
| Head and neck cancer | 33 |
| Uveal melanoma | 10 |
| LFS previvor | 29 |
| LFS survivor | 12 |
| LFS positive | 12 |
| **Baseline total** | **468** |
| *INSPIRE cohort* | |
| Breast cancer | 20 |
| Ovarian cancer | 19 |
| Head and neck cancer | 17 |
| Mixed cancer | 13 |
| Melanoma | 9 |
| **INSPIRE total** | **78** |

Table S2: **Hyperparameter search space and selected PANGEM configuration.** The identity-aware gated implementations used during model development evaluated the same candidate values and are therefore reported as a single model. Entries containing one value were held fixed during the search; entries containing multiple values were compared by internal cross-validation.

| Component | Hyperparameter | Values evaluated | Selected value |
|---|---|---|---|
| *Graph construction* | | | |
| Static scaffold | Candidate neighbours per node | $\{10, 15, 30\}$ | 15 |
| Dynamic graph | Retained neighbours per node | $\{5, 8, 10\}$ | 8 |
| Graph fusion | Static/dynamic weights | $\{0, 0.3, 0.5, 0.7, 1\}$ | 0.7/0.3 |
| *Architecture and branch fusion* | | | |
| Graph branch | Hidden-state dimension | $\{16, 32, 64\}$ | 64 |
| Graph branch | Message-passing layers | $\{1, 2, 3\}$ | 1 |
| Graph branch | Dropout probability | $\{0.1, 0.2, 0.3, 0.5\}$ | 0.5 |
| Identity embedding | Embedding dimension, $d_{\mathrm{id}}$ | $\{8, 16, 32\}$ | 16 |
| Branch fusion | Direct-branch weight | $\{0.05, 0.1, 0.15, 0.25, 0.5, 1.0\}$ | 0.1 |
| Branch fusion | Graph-branch weight | $\{1.0\}$ | 1.0 |
| Direct branch | Pooling operation | {`add`, `mean`} | `add` |
| Direct branch | Weight-initialization SD | $\{0.0001\}$ | 0.0001 |
| *Optimization during internal cross-validation* | | | |
| Optimization | Learning rate | $\{0.001, 0.0005, 0.0003\}$ | 0.0005 |
| Optimization | Weight decay | $\{0.001, 0.0005, 0.0003\}$ | 0.0005 |
| Training | Batch size | $\{16, 32\}$ | 32 |

Table S3: **Ablation study.** Graph-only variants exclude the direct DMR branch; hybrid graphs combine static and dynamic edge information, static-only and dynamic-only graphs retain the indicated edge type. Values are mean ± standard deviation across five held-out splits; multiclass metrics are macro-averaged.

| Variant | Binary AUROC | Binary AUPR | Macro-AUROC | Macro-AUPR |
|---|---|---|---|---|
| Combined direct and hybrid graph | $0.997 \pm 0.002$ | $1.000 \pm < 0.001$ | $0.977 \pm 0.027$ | $0.870 \pm 0.086$ |
| Hybrid graph only | $0.944 \pm 0.041$ | $0.991 \pm 0.007$ | $0.726 \pm 0.024$ | $0.335 \pm 0.027$ |
| Static graph only | $0.859 \pm 0.057$ | $0.976 \pm 0.010$ | $0.716 \pm 0.027$ | $0.332 \pm 0.022$ |
| Dynamic graph only | $0.915 \pm 0.037$ | $0.986 \pm 0.007$ | $0.717 \pm 0.018$ | $0.325 \pm 0.017$ |

**Effect of sample-level auxiliary features.** To assess whether complementary cfDNA measurements improved the primary methylation-based model, we compared PANGEM with an auxiliary-feature extension that added fragmentomic, fragment-ratio, nucleosome-distance, and motif-proportion profiles on the same five held-out splits. Auxiliary features did not improve mean performance across tasks (Supplementary Table S4). Binary performance was unchanged at the numerical precision. In the multiclass task, primary PANGEM had a slightly higher mean macro-AUROC and a higher mean macro-AUPR than the auxiliary-feature extension. These results indicate that the auxiliary profiles did not provide additional benefit under our framework.

Table S4: **Matched comparison of primary PANGEM and its auxiliary-feature extension.** Each seed defines a repeated stratified 90%/10% development–holdout split. Both settings used hybrid methylation graphs, the selected task-specific training protocol, and identical split assignments. The auxiliary extension added fragmentomic, fragment-ratio, nucleosome-distance, and motif-proportion encoders. Multiclass values are macro-averaged one-versus-rest metrics. Summary values are mean ± sample standard deviation across the five held-out splits.

| Task | Feature setting | Metric | Seed 42 | Seed 43 | Seed 44 | Seed 45 | Seed 46 | Mean ± SD |
|---|---|---|---|---|---|---|---|---|
| Binary | PANGEM | AUROC | 0.996 | 0.996 | 1.000 | 0.996 | 1.000 | $0.997 \pm 0.003$ |
| Binary | PANGEM + auxiliary | AUROC | 0.996 | 0.996 | 1.000 | 0.996 | 1.000 | $0.997 \pm 0.003$ |
| Binary | PANGEM | AUPR | 0.999 | 0.999 | 1.000 | 0.999 | 1.000 | $1.000 \pm < 0.001$ |
| Binary | PANGEM + auxiliary | AUPR | 0.999 | 0.999 | 1.000 | 0.999 | 1.000 | $1.000 \pm < 0.001$ |
| Multiclass | PANGEM | Macro-AUROC | 0.994 | 0.931 | 0.976 | 0.996 | 0.988 | $0.977 \pm 0.027$ |
| Multiclass | PANGEM + auxiliary | Macro-AUROC | 0.976 | 0.940 | 0.979 | 0.989 | 0.988 | $0.974 \pm 0.020$ |
| Multiclass | PANGEM | Macro-AUPR | 0.923 | 0.788 | 0.810 | 0.994 | 0.837 | $0.870 \pm 0.086$ |
| Multiclass | PANGEM + auxiliary | Macro-AUPR | 0.876 | 0.761 | 0.813 | 0.925 | 0.836 | $0.842 \pm 0.062$ |
